# Language systems from lesion-symptom mapping in aphasia: A meta-analysis of voxel-based lesion mapping studies

**DOI:** 10.1101/2021.06.03.21258096

**Authors:** Yoonhye Na, JeYoung Jung, Christopher Tench, Dorothee P. Auer, Sung-Bom Pyun

## Abstract

**Background:** Aphasia is one of the most common causes of post-stroke disabilities. As the symptoms and impact of post-stroke aphasia are heterogeneous, it is important to understand how topographical lesion heterogeneity in patients with aphasia is associated with different domains of language impairments. Here, we aim to provide a comprehensive overview of neuroanatomical basis in post-stroke aphasia through coordinate based meta-analysis of voxel-based lesion-symptom mapping studies.

**Methods:** We performed a meta-analysis of lesion-symptom mapping studies in post-stroke aphasia. We obtained coordinate-based functional neuroimaging data for 2,007 individuals with aphasia from 25 studies that met predefined inclusion criteria.

**Results:** Overall, our results revealed that the distinctive patterns of lesions in aphasia are associated with different language functions and tasks. Damage to the insular-motor areas impaired speech with preserved comprehension and a similar pattern was observed when the lesion covered the insular-motor and inferior parietal lobule. Lesions in the frontal area severely impaired speaking with relatively good comprehension. The repetition-selective deficits only arise from lesions involving the posterior superior temporal gyrus. Damage in the anterior-to-posterior temporal cortex was associated with semantic deficits.

**Conclusion:** The association patterns of lesion topography and specific language deficits provide key insights into the specific underlying language pathways. Our meta-analysis results strongly support the dual pathway model of language processing, capturing the link between the different symptom complexes of aphasias and the different underlying location of damage.

## 1. Introduction

Stroke is the most common cerebrovascular disease which occurs from interrupted or reduced blood supply or haemorrhage from the vessels to the brain tissue. The prevalence of stroke as well as stroke-related death and disability have increased over the last 25 years (Benjamin et al., 2019; Feigin et al., 2019; Lindsay et al., 2019). People suffer from various kinds of functional deficits after a stroke such as language disorders (e.g., aphasia or dyslexia), which affects at least one third of people with stroke, causing severe impact on daily life and communication. Post-stroke aphasia is heterogeneous and was classically defined by various categorical subtypes (e.g., motor [Broca] aphasia or sensory [Wernicke] aphasia) with distinct differences in the underlying lesion location identified in small autopsy studies. To better understand the granular brain-behaviour relationship in stroke populations and predict stroke outcome access to vast amount of imaging data has been instrumental (Hope, Seghier, Leff, & Price, 2013; Seghier et al., 2016). Here, we aim to provide an overview of neuroanatomical basis in post-stroke aphasia through coordinate based meta-analysis (CBMA) of voxel-based lesion-symptom mapping studies.

Over a century, lesion-symptom mapping studies have established the relationship between brain and behaviour building on the pioneering discovery by Paul Broca based on autopsy findings in a stroke patient. Traditional lesion-symptom mapping approaches have used lesion overlap and lesion subtraction methods, comparing a group of patients with a healthy or other patient control group (Damasio, 1989). More recent approaches have introduced advanced statistical methods for detailed topographical anlaysis of the brain function-lesion interrelation such as voxel-based lesion-symptom mapping (VLSM) (Bates et al., 2003), that has been widely used in stroke lesion analysis. VLSM is an image analysis technique to investigate the relationship between brain tissue damage and associated symptom development on a voxel-by-voxel basis, typically using magnetic resonance images. The first VLSM study revealed that speech fluency and language comprehension in patients with aphasia showed distinctive representations in anterior and posterior regions in the left hemisphere such that fluency was most impaired when stroke lesions affected the insular and arcuate fasciculus, and comprehension was most affected by lesions in the middle temporal gyrus (MTG) (Bates et al., 2003). Since then, VLSM has been used in examining various aspects of language impairments in post-stroke aphasia including naming (Baldo, Arévalo, Patterson, & Dronkers, 2013), reading (Piras & Marangolo, 2009), repetition (Baldo, Katseff, & Dronkers, 2012) and semantic processing (Harvey & Schnur, 2015; Schwartz et al., 2009). These findings have provided novel insights into the neuroanatomical basis of aphasia by examining the relatioship between the lesions and heterogeneous symptoms exploiting the power of automated topographical anlaysis of large and well phenotyped patient groups.

The traditional neurological model of language illustrates a division between production and comprehension within the left perisylvian cortex including Broca’s area (Berker, Berker, & Smith, 1986) and Wernicke’s area (Bogen & Bogen, 1976): lesions in Broca’s area (inferior frontal regions) disturb speech production, lesions in Wernicke’s area (posterior superior temporal regions) interrupt language comprehension, and the disconnection between these regions (lesions in arcuate fasciculus) disrupts their communication, resulting in the failure in repetition of heard speech (Lichtheim, 1885). Building on rich evidence from neurological and functional imaging studies, Hickock and Poeppel (Hickok & Poeppel, 2000, 2004) and others proposed a dual stream model that language is subserved by two distinctive pathways (Warren, Wise, & Warren, 2005; Wise, 2003). The ventral stream projects ventrolaterally to middle and inferior posterior temporal cortex which serves as an interface between sound-based representations of speech to widely distributed conceptual representations. The dorsal stream involved in mapping sound onto articulatory representations projects dorsoposteriorly toward inferior parietal and frontal regions. Most recent models include brain areas beyond the classical perisylvian language areas such as the anterior temporal lobe involved in processing of word and sentence meaning (Binder & Desai, 2011; Dronkers, Wilkins, Van Valin, Redfern, & Jaeger, 2004; Patterson, Nestor, & Rogers, 2007). In addition, the advance of diffusion tensor imaging has led to significant understanding of white matter pathways in the brain and language, providing re-evaluation of classical disconnection accounts for aphasia. Yet, there is limited consensus and granularity of functional anatomical associations of specific language deficits from stroke patients. Accordingly we aim to generate overall understandings between the lesions and language processing in post-stroke aphasia making use of advanced spatial meta-analysis of VSLM studies from large populations.

In this study, we investigated the neuroanatomical baisis of lanugage from post-stroke aphasia, drawing on a meta-analysis of published lesion-symptom mapping studies. Our work evaluated the concurrence between findings from various lesion-symtopm mapping studies by employing coordinate based meta-analysis. Specifically, we examined (i) the overall convergnece between the results from studies concerned language comprehension and production, (ii) the concurrence in lesion sites associated with various language function such as semantics, phonology, and speech fluency, and (iii) the concurrence in lesion sites associated with deficits in language tasks such as repetition, naming, and reading. Our analyses evalutated whether bahavioural differences in language processing can be matched with different lesion patterns in stroke patients. We expect that language deficits in post-stroke aphasia link to both distinct and common patterns of lesions. Our results would provide insights into the discrepancies that exist in previous literaure examining lesions in post-stroke aphasia. We will discuss our findings in terms of functional accounts of aphasia and theoretical language models based on both healthy and impaired language functions.

## 2. Methods

### 2.1 Literature search and selection

We undertook a literature search of scopus (http://www.scopus.com), pubmed (http://pubmed.ncbi.nlm.nih.gov) and google scholar (http://scholar.google.com) databases for lesion mapping and/or VLSM studies with stroke patients published in English. Searching keywords were the combinations of several terms from various language functions: ‘stroke’, ‘cerebral infarction’, ‘cerebral hemorrhage’, ‘aphasia’, ‘post-stroke’, ‘language’, ‘comprehension’, ‘production’, ‘speech’, ‘semantic’, ‘word’, ‘lexical’, ‘naming’, ‘repetition’, ‘sentence’, ‘phonology’, ‘syntactic’, ‘phonotactic’, ‘grammar’, ‘voxel’, ‘voxel-based’, ‘VLSM’, ‘lesion’ and ‘lesion mapping’.

Two researchers (JJ and YN) independently conducted the literature search, and reviewed the methodological quality of the assessed behavioural tests of the original studies. Finally, 568 studies were found from the databases with searching time as “all year” till April, 2020, and overlapping articles were removed. One hundred six relevant studies were remained after the abstract screening and 58 articles were removed after full paper reading. Finally, 25 articles were entered into the meta-analysis (Table 1). The final 25 articles were selected out of 48 candidates, following the inclusion criteria: i) studies reported brain coordinates of the lesion mapping results in a standard space (MNI or Talairach), ii) studies using neuropsychological batteries (e.g., Philadelphia Naming Test (Roach, Schwartz, Martin, Grewal, & Brecher, 1996), Comprehensive Aphasia Test (Swinburn, Howard, & Porter, 2004), The Western Aphasia Battery (Risser & Spreen, 1985), and etc.) assessing language functions and/or language task, iii) studies conducting whole brain analysis. Figure 1 illustrates the process of study search and selection. And the exclusion criteria were as follows: i) studies unrelated to the language function, ii) studies conducted without neuroimaging (e.g., neurosurgical research), iii) studies reported results without the brain coordinates.

**Table 1.**
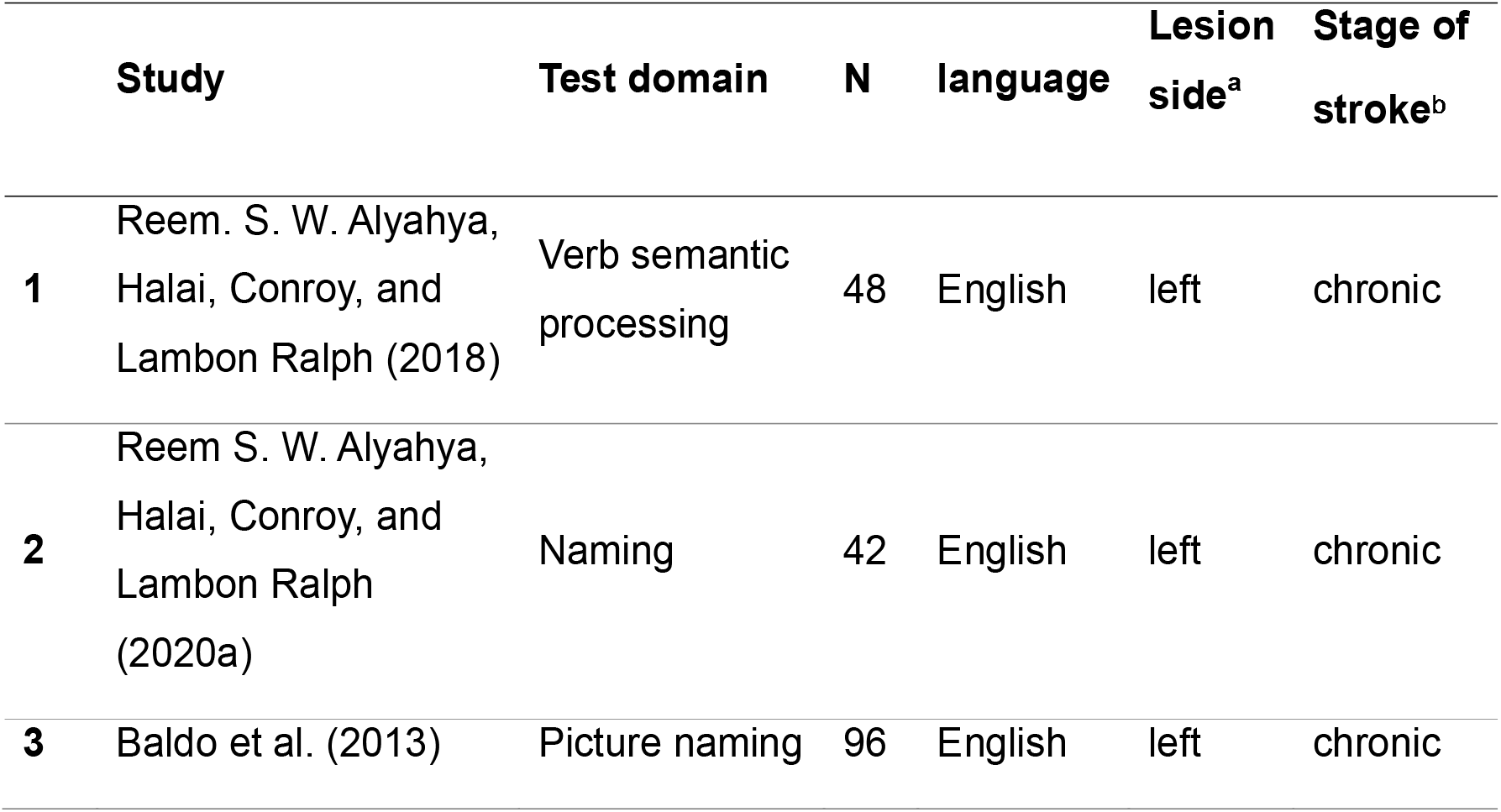
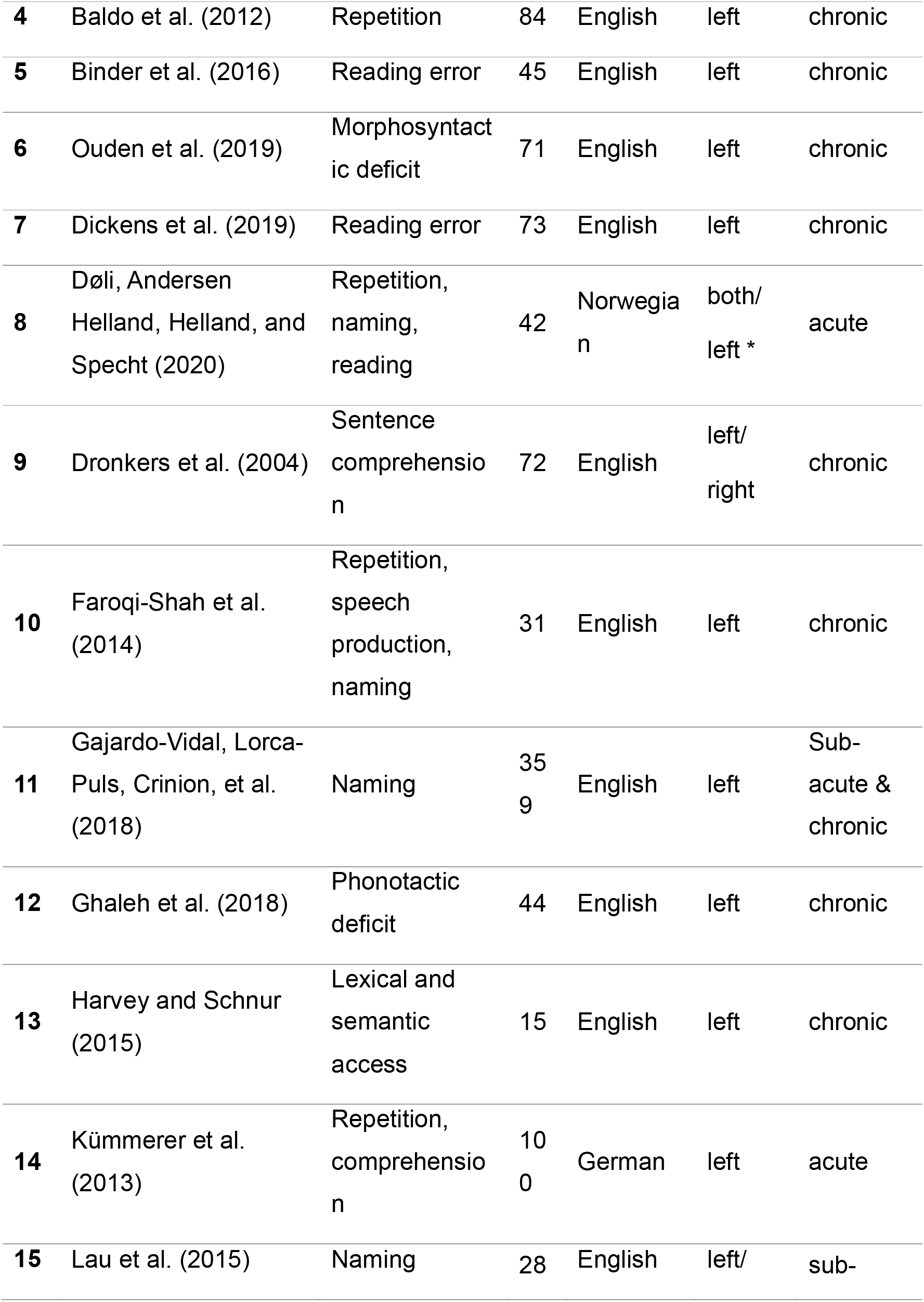
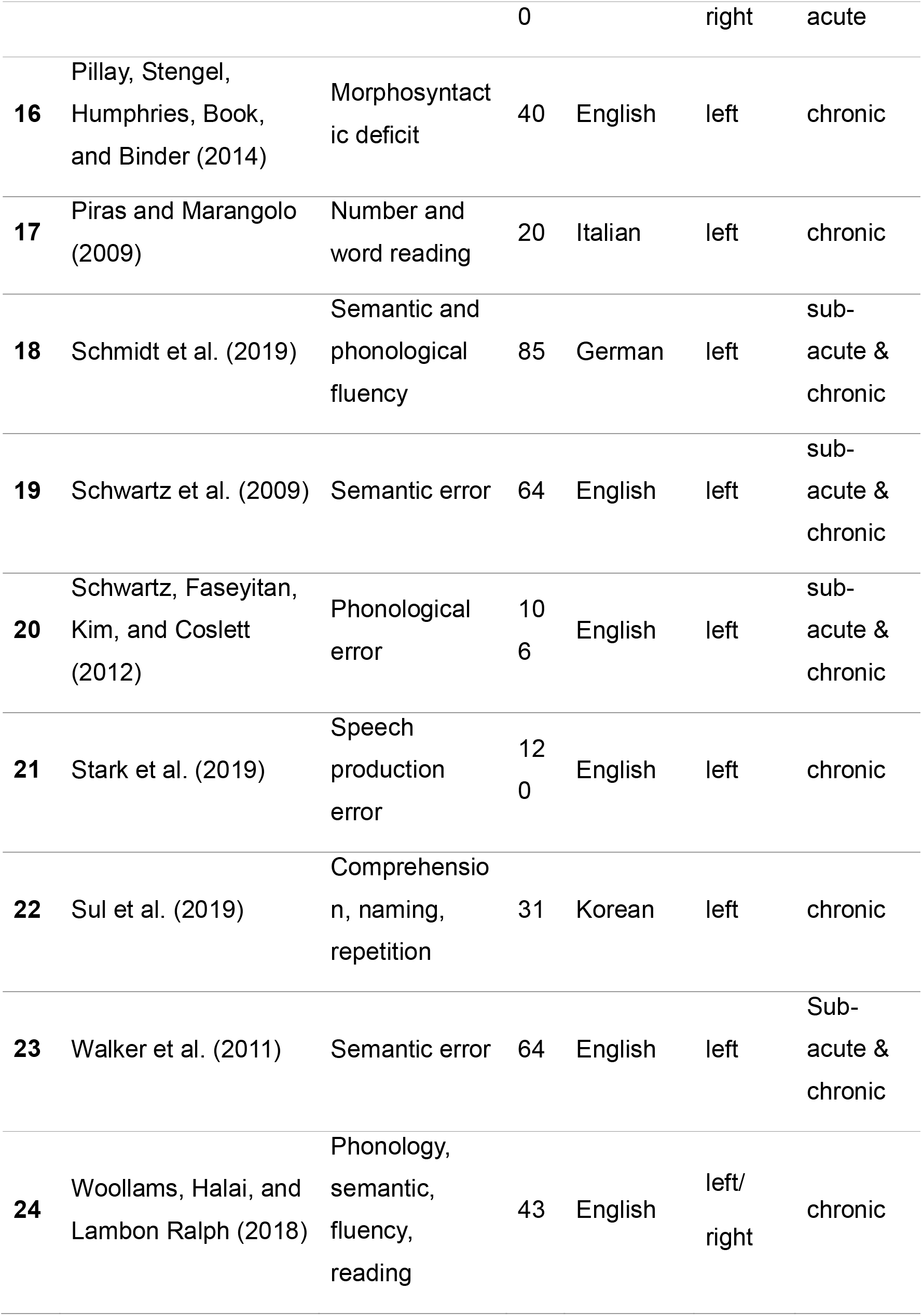
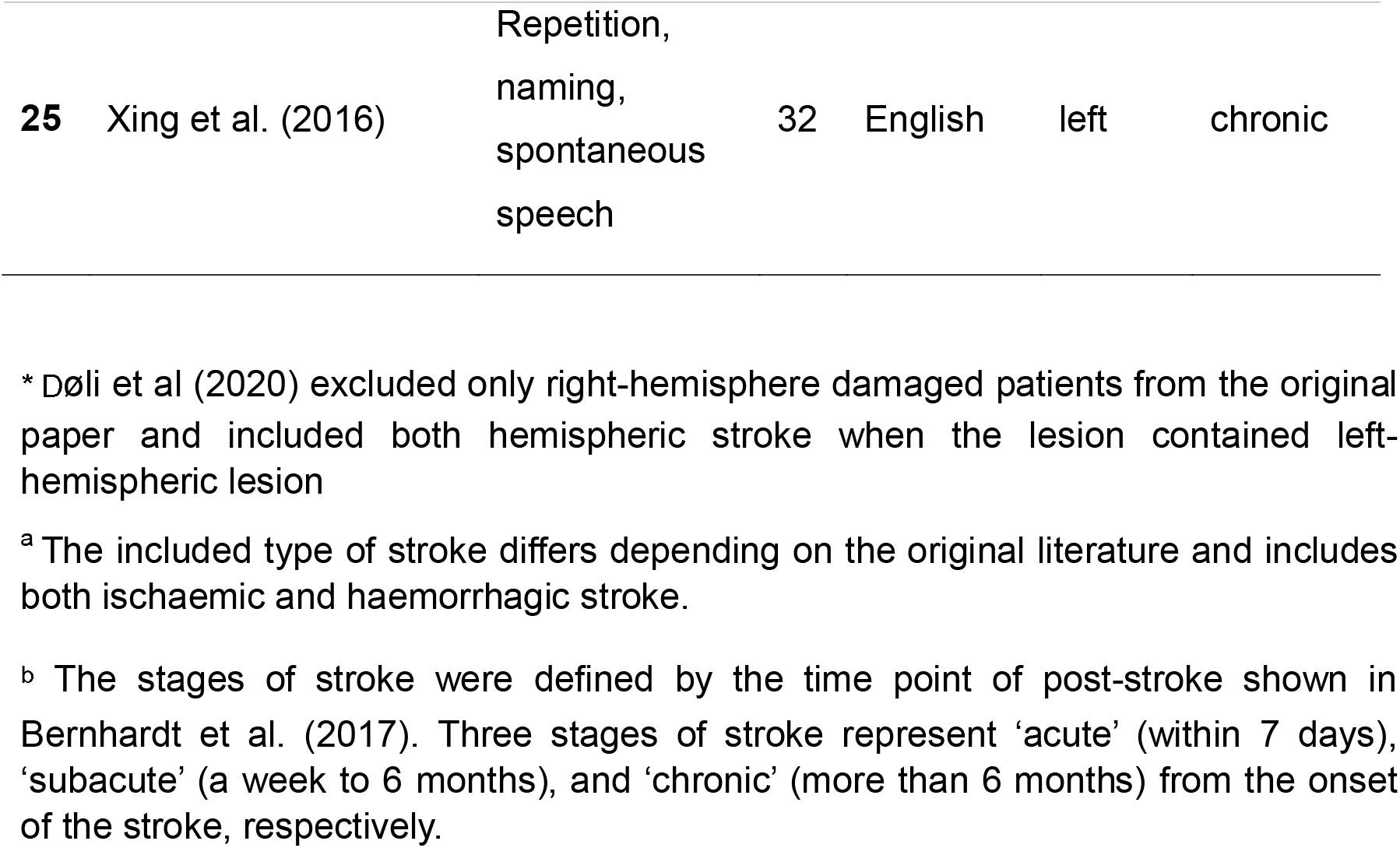
The list of 25 studies included in the meta-analysis.

**Figure 1.**
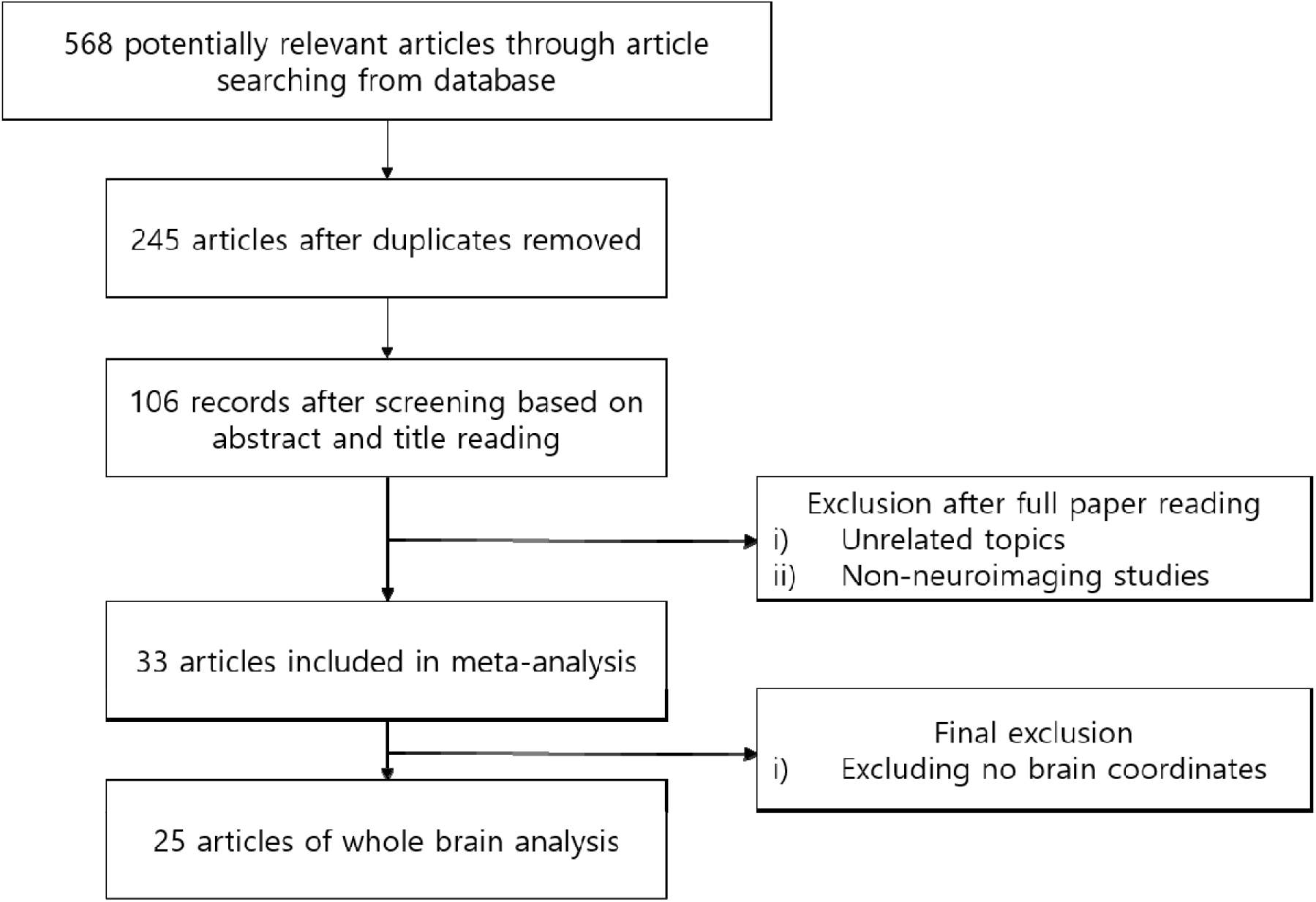
Flowchart of lesion-mapping studied included in the analysis

### 2.2 Data analysis

In order to examine the common and distinctive lesion patterns in relation to language deficits in aphasia, we performed several different meta-analyses (Table 2). The first analysis included all data from all papers reporting the neural substrates of aphasia, providing the overall lesion patterns associated with aphasia. Then, we examined the link between language functions and lesion patterns, categorizing the neuropsychological batteries/tasks into language comprehension and production, respectively. Comprehension category included studies tested language receptive function such as verb comprehension or word-picture matching, whereas production category comprised of studies tested overall expressive language function such as speech fluency, sentence production and word/number reading aloud. Then, we categorized the data for a specific language function such as semantic or phonology when they meet the sufficient number of studies more than 3. Accordingly, semantics, phonology and speech fluency were included in the meta-analysis. Semantics included studies used sematic fluency or semantic error, and phonology contained studies tested phonological fluency or phonological error/deficits. Speech fluency category comprised of studies tested spontaneous speech fluency/frequency or sentence production from picture description. Additional analyses were conducted for task types including repetition, naming and reading.

**Table 2.** the number of studies, foci, and description or examples of the tasks according to language functions/tasks

| Category | Description/examples | N of study | N of foci |
| --- | --- | --- | --- |
| **General language** | overall language function | 25 | 424 |
| **Comprehension** | sentence comprehension, word-picture matching | 5 | 84 |
| **Production** | sentence production, fluency, speech, naming, reading aloud, reading error | 20 | 313 |
| **Semantics** | semantic error, semantic fluency | 16 | 144 |
| **Phonology** | phonological fluency, phonological error | 13 | 128 |
| **speech fluency** | spontaneous speech, sentence production, naming frequency | 4 | 44 |
| **Repetition** | word or non-word repetition | 6 | 29 |
| **Naming** | verb naming, Philadelphia naming test, naming error | 12 | 92 |
| **Reading** | word reading, number reading, reading error | 5 | 73 |

### 2.3 Coordinate based meta-analysis (CBMA)

We performed the meta-analyses using LocalALE tool implemented in NeuRoi (https://www.nottingham.ac.uk/research/groups/clinicalneurology/neuroi.aspx) (Tench, Tanasescu, Auer, & Constantinescu, 2013). Extracted lesion coordinates were entered into a series of CBMA to estimate the concurrence between the reported lesions in aphasia from different published studies and to examine common and different lesion patterns underlying different language deficits in aphasia. The detailed description of the method for generating the likelihood estimates can be found in Tench et al. (2013). LocalALE models each reported coordinate foci as a spatial 3D truncated Gaussian distribution and is similar to the popular activation likelihood estimation (ALE) method (Eickhoff et al., 2009; Turkeltaub, Eden, Jones, & Zeffiro, 2002), but modified to overcome some limitations. Importantly, the full width half maxima (FWHM) was automatically modified to account for the number of studies included in the analysis helping to prevent false positive results for large number of studies and false negatives for small number of studies (Tench, Tanasescu, Auer, Cottam, & Constantinescu, 2014). Furthermore, LocalALE does not require a minimum of at least 17 studies to avoid individual studies dominating the significant results (Eickhoff et al., 2016). The results were considered significant by cluster-wise thresholding at *p* < 0.05 from a nonparametric permutation test after false cluster discovery rate (FCDR) correction (Tench et al., 2013); this performs a correction such that the proportion of clusters expected to be significant under the null hypothesis of random coordinates is only 5%. All results are reported in Talairach space.

### 2.4 Neurosynth

In order to link the lesion sites identified from VSLM meta-analyses to functional systems derived from published brain activation studies, we performed additional meta-analyses using Neurosynth (http://neurosynth.org). Neurosynth is an online platform offering automated synthesis of functional MRI data on the basis of large-scale database (Yarkoni, Poldrack, Nichols, Van Essen, & Wager, 2011). We used the peak coordinate of each cluster from our CBMA results as a seed region (a 6mm sphere) to generate the meta-analytic coactivation maps and keywords associated with the maps. All the coordinates were converted from Talairach system to MNI coordinates and entered into Neurosynth. Neurosynth produces a z-score map that corresponds to the likelihood that a term of interest had been used in a journal article given the presence of activation. The Neurosynth results were voxel-wise thresholded at the level of 0.01 after FDR correction. Keywords were extracted on the basis of the z-score was greater than 0 and up to 10 keywords were chosen in order of the z-score.

## 3. Results

### 3.1 CBMA analyses

Twenty-five studies (2,007 aphasia patients; total of 45 experiments with 424 relevant foci; 410 in the left hemisphere and 14 in the right) that met the inclusion criteria were identified and their data were entered into the analyses. The results are summarized in Figure 2 and Table 3. The primary analysis with all data revealed the regions within the left perisylvian cortex including the middle and superior temporal gyrus (MTG and STG, respectively), inferior frontal gyrus (IFG), insula, precentral gyrus, fusiform gyrus (FG), and inferior parietal lobule (IPL) (Fig. 2a). And there was no significant cluster in the right hemisphere due to small number of input foci. The analyses examining the language comprehension and production showed the distinctive patterns for each function: two significant clusters in the STG and FG associated with comprehension impairments (Fig. 2b) and six significant clusters associated with language production deficits including the IFG, precentral gyrus, STG, MTG, and insula (Fig. 2c).

**Table 3.**
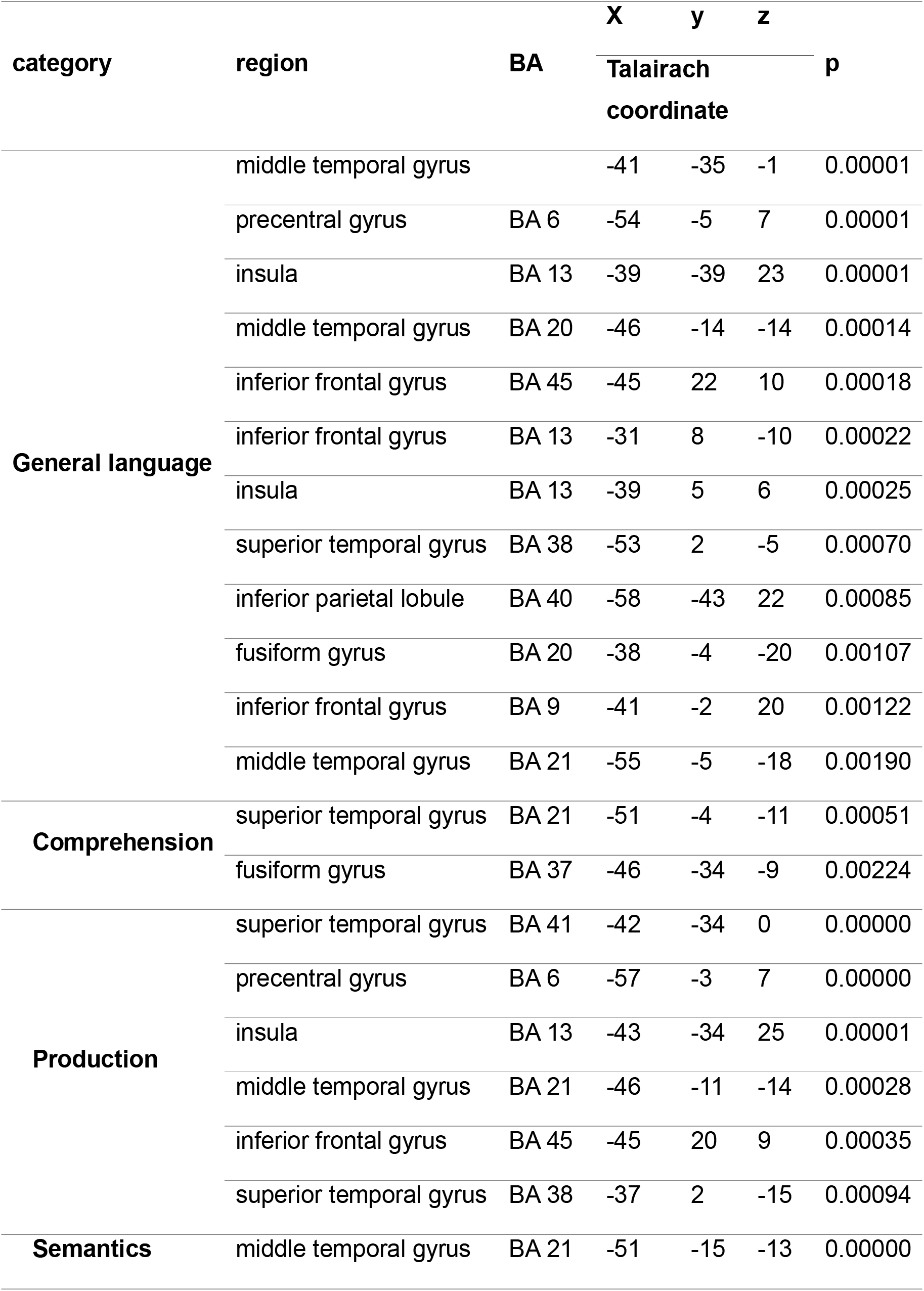
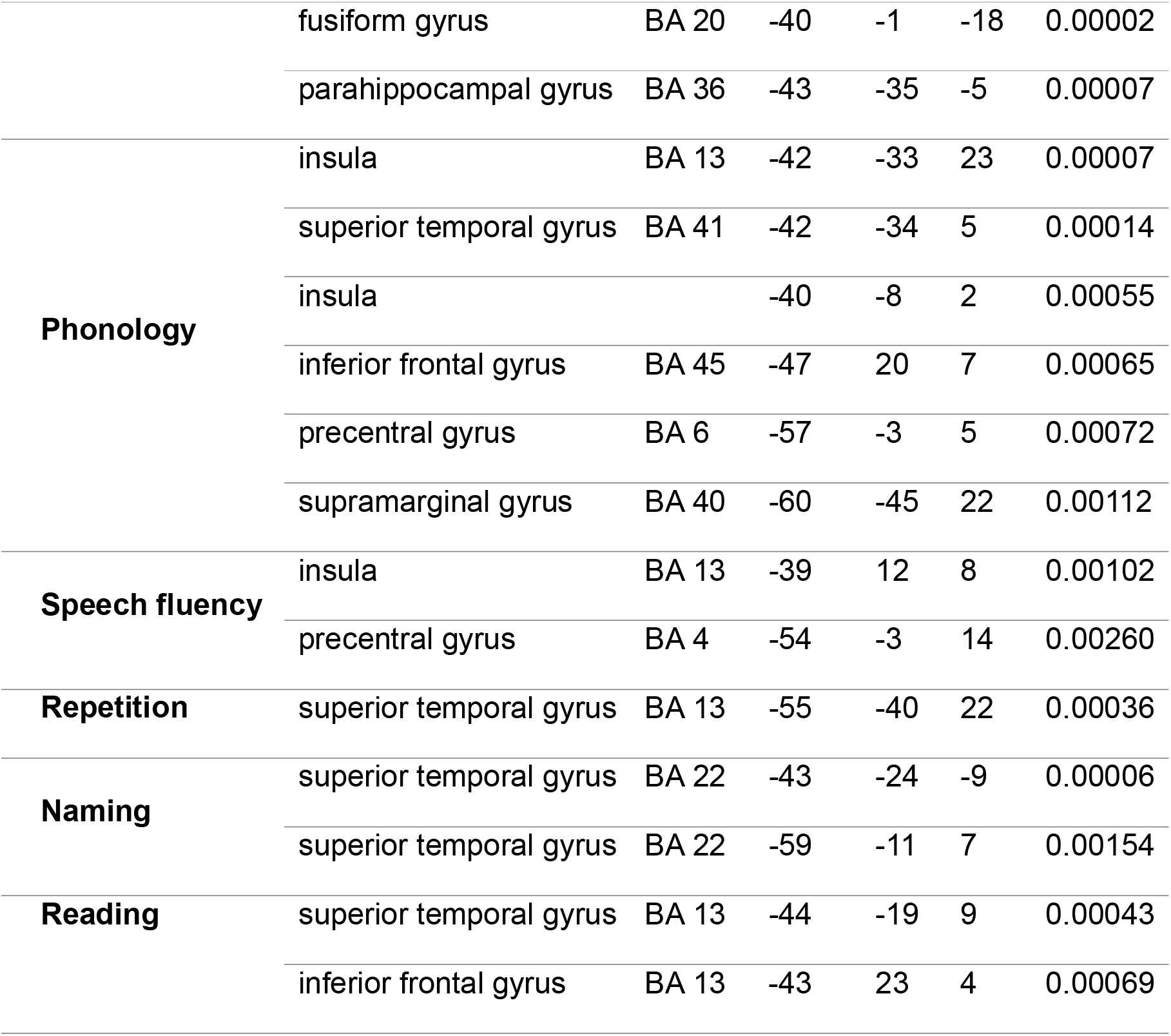
the summary of the meta-analyses. BA = Brodmann’s area

**Figure 2.**
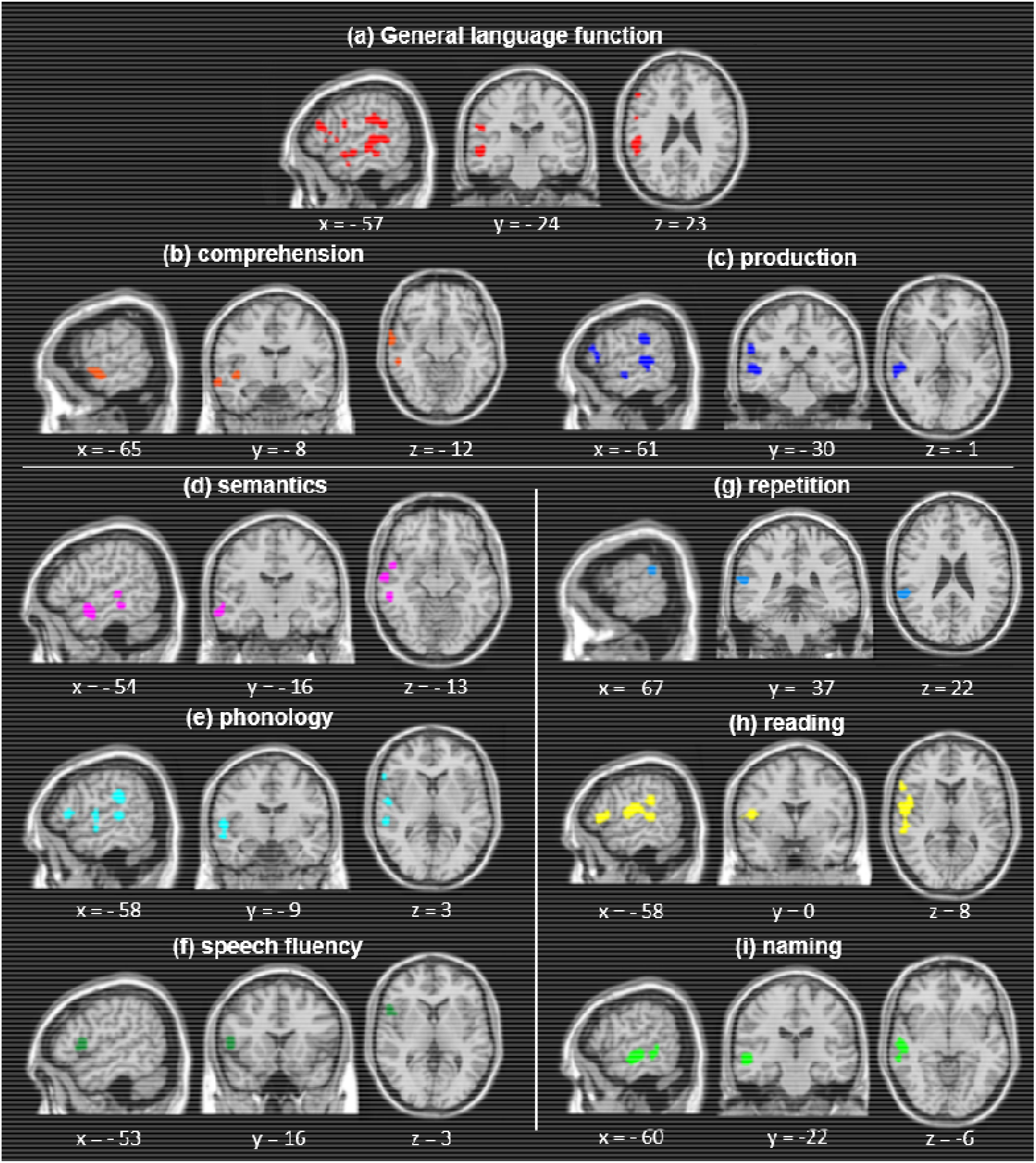
meta-analysis results. (a) result for the general language function. (b-f) results according to language functions. (g-i) task-based results.

Distinctive lesion patterns were found according to language functions. Semantic function was associated with anterior and middle MTG, FG, and parahippocampal gyrus (Fig. 2d). Phonological deficits were involved in the insular, IFG, precentral gyrus, STG, and supramarginal gyrus (SMG) (Fig. 2e). Speech fluency was associated with lesions in the insula, IFG, and precentral gyrus (Fig. 2f). We found task-specific lesion patterns in aphasia. The posterior STG was associated with repetition (Fig. 2g). Reading impairments were related to the ventral IFG, temporoparietal junction (TPJ), and posterior STG (Fig. 2h), overlapping with the phonology results (Fig. 2e). Naming task revealed the middle/posterior STG and MTG (Fig. 2i), overlapping with the results of semantic (Fig. 2d).

There were systematic differences in the patterns of lesions across language functions/tasks (Fig. 3). Within the left perisylvian cortex, the ventral regions in the temporal lobe were associated with comprehension and semantic processing, whereas the dorsal areas in frontoparietal cortex were involved in expressive language function, phonology, and speech fluency. In summary, our findings suggest the neuroanatomical basis of language deficits in aphasia, supporting the contemporary dual pathway model of language processing (Hickok & Poeppel, 2004; Saur et al., 2008; Ueno, Saito, Rogers, & Lambon Ralph, 2011).

**Figure 3.**
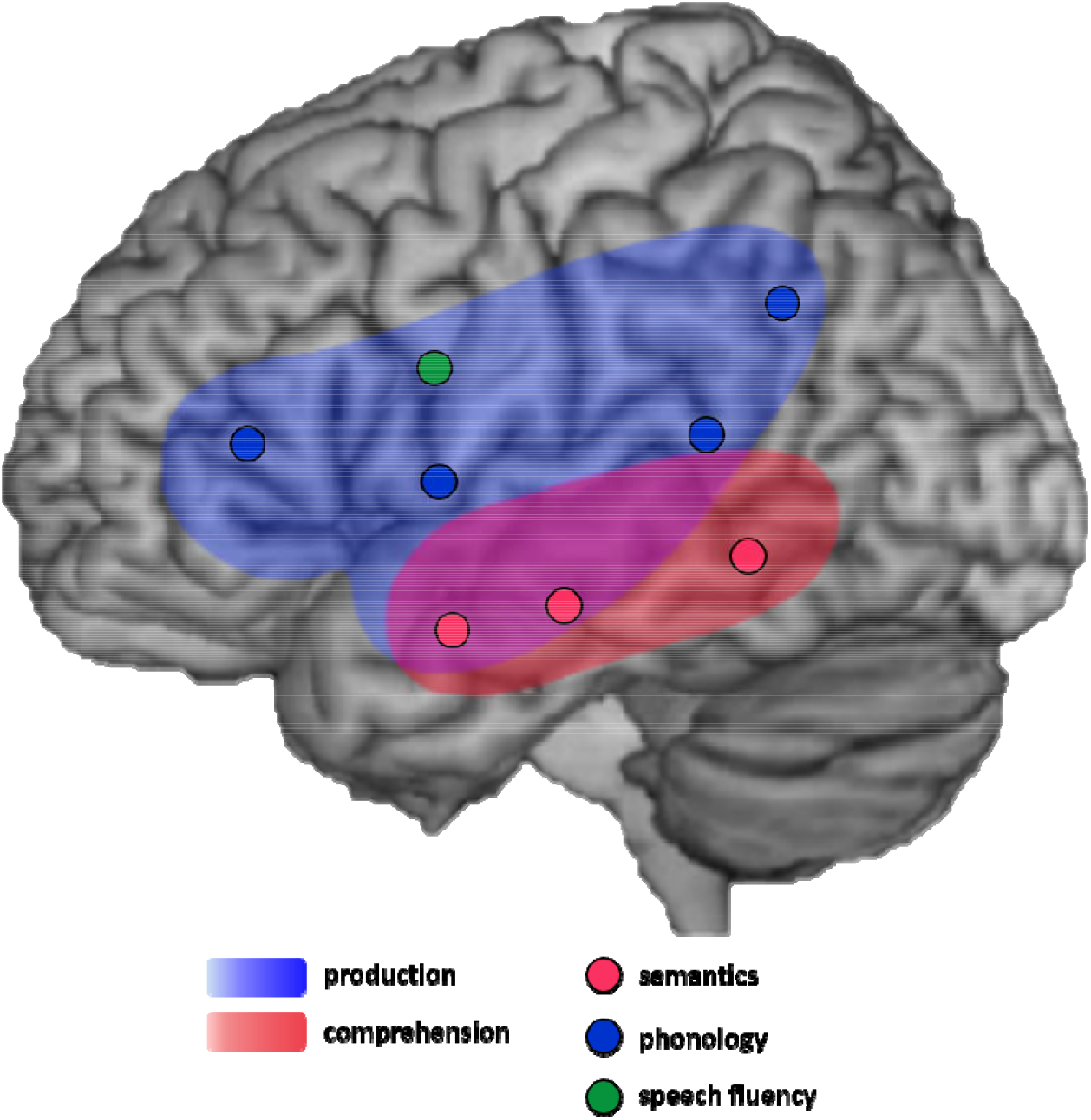
A schematic diagram summarizing the meta-analysis results. Colors (pink and sky-blue) indicate the coverage of general language functions, production and comprehension, respectively. Each dot points each peak cluster according to functional categories.

### 3.2 Neurosynth

In order to examine functional systems associated with the clusters from our meta-analysis, we performed meta-analytic coactivation analysis using Neurosynth. The results are summarized in the Figure 4 and Table S1 & S2. The coactivation maps of the semantic clusters included the IFG, medial prefrontal cortex, middle cingulate cortex (MCC), STG, MTG, SMG, AG, thalamus, putamen, and caudate (Figure 4a). The related keywords of the coactivation maps of semantic clusters were ‘semantic’, ‘read’, ‘word’, and ‘comprehension’. The clusters of the phonology were coactivated with the IFG, MTG, and insular (Figure 4b) and the related keywords were ‘auditory’, ‘speech’, ‘listening’, and ‘sound’. The coactivation maps of speech fluency clusters covered the insular, MCC, supplementary motor area (SMA), thalamus, cerebellum, and visual cortex (Figure 4c) and the related keywords were ‘phonological’, ‘auditory’, ‘speech perception/production’, and ‘acoustic’.

**Figure 4.**
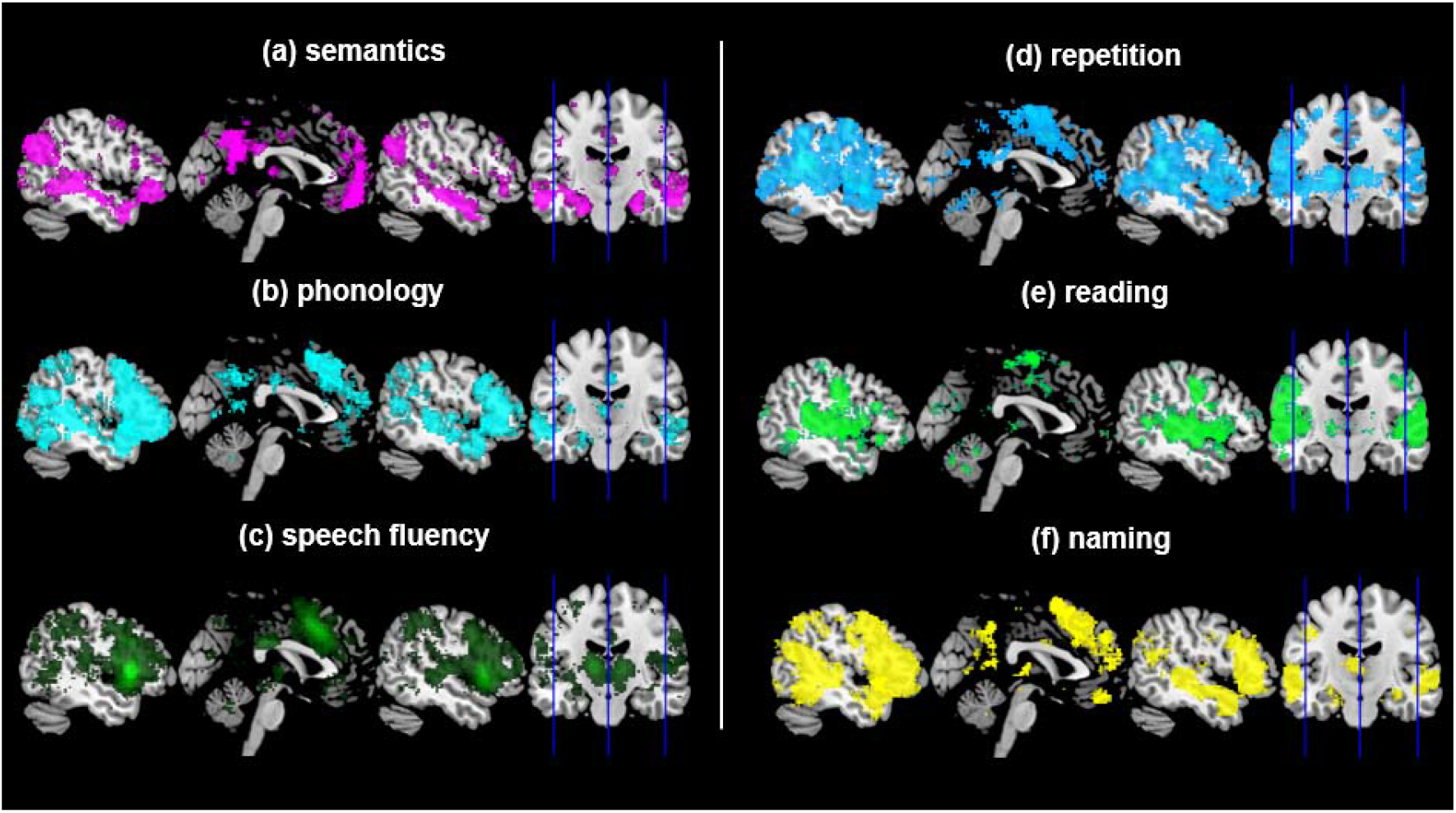
The meta-analytic coactivation maps from the Neurosynth. Each figure represents the functionally coactivated region with the peak cluster of the CBMA results. Each sagittal plane is shown at x=-50, 0, 50.

The task-related clusters revealed the similar patterns of meta-analytic coactivated maps covering the fronto-temporo-parietal system. The repetition clusters were coactivated with the IFG, MCC, STG, SMG, superior parietal lobe (SPL), and precuneus (Figure 4d) and involved in keywords such as ‘auditory’, ‘motor’, ‘hearing’, ‘production’, ‘phonological’, and ‘mirror’. The reading clusters revealed the coactivation maps including the IFG, STG, MTG, IMG, FG, SMG, AG, cerebellum, and visual cortex (Figure 4e) and the related keywords were ‘auditory’. ‘listening’, ‘language’, ‘comprehension’, ‘semantic’, ‘words’, and ‘sentences’. The naming clusters were coactivated with the IFG, MCC, SMA, rolandic operculum, STG, MTG, ITG, FG, hippocampus, parahippocampal gyrus, posterior cingulate cortex, precuneus, cerebellum, thalamus, caudate, and visual cortex (Figure 4f). These maps were associated with keywords such as ‘episodic’, ‘recall’, amnestic’, ‘semantic memory’, ‘auditory’, and ‘speech’.

## 4. Discussion

In order to identify the lesion patterns in post-stroke aphasia and to investigate whether there are consistent differences in the patterns of lesions across different language function and tasks, we performed a coordinate based meta-analysis of lesion-symptom mapping studies in post-stroke aphasia. We obtained coordinate-based lesion-symptom mapping data from 2,007 aphasia patients and identified different patterns of function- and task-specific lesions associated with language processing. Using the additional meta-analytic coactivation approach with the clusters, we identified the functional neural systems associated in healthy controls with the lesion sites identified in post-stroke aphasia. The CBMA results revealed that the overall patterns of the language processing such as the production and the comprehension was in line with the dual pathway model (Hickok & Poeppel, 2004; Ueno et al., 2011) and a specific pattern of lesion in the model was associated with selective deficits in language functions. Especially, damage to the frontal lobe impaired language production including phonology and speech fluency and lesions in the dorsal-posterior temporal lobe was exclusively associated with the repetition task. In contrary, the lateral temporal lobe lesions were attributed to impairment in language comprehension including semantic processing. The results provide novel insights into the neuroanatomical basis of lanugage processing from post-stroke aphasia.

The patterns of lesion sites in post-stroke aphasia were broadly consistent with dual pathway models of language processing that propose a dorsal pathway connecting frontal and temporal-parietal cortex for language production and a ventral pathway linking temporal-frontal cortex for language comprehension (Hickok & Poeppel, 2000; Saur et al., 2008; Scott, 2000; Ueno et al., 2011). According to the dual-pathway model (Hickok & Poeppel, 2004), the dorsal route serves speech production as well as auditory-motor integration, the essential function for speech production and some speech recognition tasks. Our results supports the model demonstrating that patients with post-stroke aphasia had language production deficits with lesions in the dorsal route connecting from the supramarginal gyrus through precentral sensorimotor regions and insular involved in articulatory motor control to the inferior frontal cortex (Reem S. W. Alyahya, Halai, Conroy, & Lambon Ralph, 2020b; McKinnon et al., 2018). In the present study, the lesion sites of language production overlap with the ventral pathway including the superior and middle temporal gyrus. A meta-analysis of functional neuroimaging studies reported that the auditory ventral stream covering the anterior-to-posterior STG were associated with the speech processing from recognition of speech sounds to spoken words (Dewitt & Rauschecker, 2012). Our results converge with previous findings that language production accompanies the intrinsic comprehension of words or objects used in the task (M. A. Lambon Ralph, McClelland, Patterson, Galton, & Hodges, 2001; Morton, 1985) and that language production tasks such as reading aloud and naming are attributed to the ventral pathway (Moore & Price, 1999).

Subsequent analyses for a specific language function and task in production revealed the similar pattern of lesions. The CBMA results of phonology and reading were identical to the lesions of language production including the IFG, precentral gyrus, insula, SMG and STG. fMRI studies have demonstrated that these regions are associated with phonological processing in healthy individuals (Burton, Small, & Blumstein, 2000; Gold, Balota, Kirchhoff, & Buckner, 2005; Poldrack et al., 2001). Especially, the STG is involved in phonological aspects of both speech perception and production (Buchsbaum, Hickok, & Humphries, 2001; Burton, Small, & Blumstein, 2000; Gold, Balota, Kirchhoff, & Buckner, 2005; Poldrack et al., 2001). Importantly, these regions in the dorsal pathway are connected via arcuate fasciculus which plays a crucial role in conducting phonological and motor processing of articulation for speaking (Catani & Mesulam, 2008; Saur et al., 2008). Our analysis for speech fluency revealed two clusters in the insula and precentral gyrus. These regions have been repeatedly associated with articulatory planning (Basilakos, Smith, Fillmore, Fridriksson, & Fedorenko, 2018; Dronkers, 1996) and motor coordination of speech (Ackermann & Riecker, 2004; Baldo, Wilkins, Ogar, Willock, & Dronkers, 2011). Accumulating clinical and functional imaging evidence highlights the role of the left insula in speech motor control, supporting the temporo-spatial pattern of vocal track muscle innervation during speech (Ackermann & Riecker, 2004). Contrary to other sub-categories of language production, we found one cluster associated with repetition in the left posterior STG (pSTG). The evidence supporting the involvement of pSTG in the repetition task comes from conduction aphasia characterized by good auditory comprehension, fluent speech, and the inability to repeat words or phrases (Buchsbaum et al., 2001; Hickok et al., 2000). Furthermore, direct electrical stimulation over the pSTG induced conduction aphasia-like symptoms (Anderson et al., 1999). Thus, our data supports the view that conduction aphasia is a disorder of cortical dysfunction rather than a disconnection syndrome (Buchsbaum et al., 2001; Hickok et al., 2000). Our findings directly fit into the neuroanatomical-constrained dual-pathway models of language, showing the impact of lesions to the perisylvian regions. Damage to the insular-motor areas impaired spoken output with preserved comprehension and a similar pattern was observed when the lesion covered the insular-motor and SMG. Lesions in the IFG severely impaired speaking with relatively good comprehension. The repetition-selective deficits only arise from the lesion the pSTG (Ueno et al., 2011). Overall, our results suggest these regions in the dorsal pathway serves a crucial role in recognizing and generating speech and a damage in this pathway produces various deficits in language production function, specific to the different patterns of lesions in aphasia.

Our results demonstrated the role of the ventral stream along with the anterior-to-posterior temporal cortex in language comprehension, especially semantic processing (Hickok & Poeppel, 2000; Saur et al., 2008; Scott, 2000; Ueno et al., 2011). Specifically, the anterior temporal lobe (ATL) is a transmodal hub for semantic representation (Matthew A Lambon Ralph, Jefferies, Patterson, & Rogers, 2017) supported by converging evidence from patients’ studies (Bozeat, Lambon Ralph, Patterson, Garrard, & Hodges, 2000; Hodges & Patterson, 2007), intracranial recording (Chen et al., 2016; Shimotake et al., 2015), fMRI (Coutanche & Thompson-Schill, 2015; Peelen & Caramazza, 2012), and brain stimulation(Jung & Lambon Ralph, 2016, 2021). The computational modelling of the dual-pathway also supports that damage in the ATL (STG and MTG) generated impairment in comprehension and naming. Consistent with Ueno et al (2011)’s model, semantic fluency, naming, and naming errors were strongly associated with the anterior and lateral STG and MTG damage in post-stroke aphasia patients. As the ATL receives inputs from auditory and visual cortex, showing the graded sensory-to-semantic rostral progression (Jung, Cloutman, Binney, & Lambon Ralph, 2017; Jung, Visser, Binney, & Lambon Ralph, 2018; Scott, 2000), the neighboring STG and fusiform gyrus found in our results may be involved in the progression in semantic processing.

Despite of the large number of patients and clear findings, this study has limitations. Most of patients included had a single left hemisphere stroke, which led to no CBMA clusters in the right hemisphere. As language functions are lateralized to the left hemisphere, the most original literature reviewed in this study included only patients with left hemispheric lesion. However, recent studies have demonstrated that the right hemisphere damage after stroke cause language disorders (Gajardo-Vidal, Lorca-Puls, Hope, et al., 2018; Ross, 1981). It might explain the discrepancy between our meta-analysis results and Neurosynth findings. The functional neural systems connected to the lesion sites resided in the bilateral hemispheres including prefrontal, temporal, and parietal cortices. A recent fMRI meta-analysis in post-stroke aphasia showed bilateral activation in the fronto-temporo-parietal network during language processing in aphasic patients with the left hemisphere lesion (Stefaniak, Alyahya, & Lambon Ralph, 2020). Thus, future studies are warranted to investigate stroke patients with right hemisphere lesions to elucidate the relationship between the right hemisphere damage and language deficits in post-stroke aphasia. Finally, most languages used in the original literature were English. Future studies are required to have languages varieties in order to confirm our findings in patients with aphasia across different languages.

In conclusion, our results strongly support the dual pathway model of language processing, capturing the link between the different symptom complexes of aphasias and the different underlying location of damage. The patterns of lesions provide key insights about the underlying process of each language pathway. The present study demonstrates the functional and neural architecture of language system.

## Data Availability

The data that support the findings of this study are available on request from the corresponding author, DPA and SBP.

## Author contributions

JJ, DPA, and SBP conceptualised the study and designed the research; YN and JJ obtained and analyzed the data; YN and CT performed the statistical analysis; YN and JJ, wrote the initial manuscript. All co-authors edited the manuscript.

