## supplementary table 1&2 for "Language systems from lesion-symptom mapping in aphasia: A meta-analysis of voxel-based lesion mapping studies"

**Supplementary Information**

**Supplementary Table 1 & 2**

**Supplementary Table 1**

| cluster | Number of voxel | x | y | z | label |
| --- | --- | --- | --- | --- | --- |
| *Functional category* | | | | | |
| *semantics* | | | | | |
| cluster1 | 24918 | -54 | -16 | -22 | L Middle Temporal Gyrus |
|  |  | 58 | -12 | -18 | R Middle Temporal Gyrus |
|  |  | 62 | -12 | -18 |  |
|  |  | 58 | -18 | -18 |  |
|  |  | 54 | -2 | -20 |  |
|  |  | 58 | -4 | -18 |  |
|  |  | 58 | 0 | -20 |  |
|  |  | 50 | 2 | -24 |  |
|  |  | 56 | 2 | -24 |  |
|  |  | -48 | -66 | 28 | L Angular Gyrus |
|  |  | 52 | -8 | -14 | R Superior Temporal Gyrus |
| cluster2 | 1267 | 50 | -64 | 30 | R Angular Gyrus |
|  |  | 54 | -66 | 30 |  |
|  |  | 50 | -66 | 34 |  |
|  |  | 48 | -70 | 36 |  |
|  |  | 52 | -56 | 36 |  |
|  |  | 58 | -56 | 16 | R Middle Temporal Gyrus |
|  |  | 58 | -48 | 34 | R SupraMarginal Gyrus |
|  |  | 60 | -44 | 38 |  |
|  |  | 64 | -34 | 32 |  |
|  |  | 64 | -32 | 38 |  |
|  |  | 56 | -56 | 40 | R Inferior Parietal Lobule |
| cluster3 | 357 | 24 | 34 | 44 | R Superior Frontal Gyrus |
|  |  | 22 | 42 | 40 |  |
|  |  | 24 | 18 | 50 |  |
|  |  | 20 | 18 | 52 |  |
|  |  | 16 | 44 | 44 |  |
|  |  | 12 | 26 | 38 | R MCC |
|  |  | 24 | 28 | 38 | R Middle Frontal Gyrus |
|  |  | 30 | 30 | 46 |  |
|  |  | 26 | 34 | 40 |  |
|  |  | 24 | 24 | 36 |  |
|  |  | 26 | 14 | 46 |  |
| cluster4 | 289 | -42 | 8 | 48 | L Precentral Gyrus |
|  |  | -48 | -2 | 54 |  |
|  |  | -54 | 2 | 48 |  |
|  |  | -56 | 2 | 44 |  |
|  |  | -48 | 6 | 50 |  |
|  |  | -44 | 0 | 54 |  |
|  |  | -52 | 0 | 40 |  |
|  |  | -46 | 10 | 52 | L Middle Frontal Gyrus |
|  |  | -56 | -4 | 40 | L Postcentral Gyrus |
| cluster5 | 205 | -24 | 6 | -10 | L Putamen |
|  |  | -18 | 14 | -6 |  |
|  |  | -20 | 12 | -10 |  |
|  |  | -14 | 14 | -8 |  |
|  |  | -16 | 18 | -8 |  |
|  |  | -26 | 0 | -6 |  |
|  |  | -24 | 8 | -14 | L Olfactory cortex |
|  |  | -12 | 22 | -8 | L Caudate Nucleus |
| cluster6 | 122 | 6 | -16 | 12 | R Thalamus |
|  |  | -6 | -18 | -2 | L Thalamus |
|  |  | -4 | -18 | 2 |  |
|  |  | -54 | 2 | 48 | L Precentral Gyrus |
|  |  | -56 | 2 | 44 |  |
| *phonology* | | | | | |
| cluster1 | 28184 | -48 | 20 | -4 | L IFG (p. Orbitalis) |
|  |  | -54 | 22 | 20 | L IFG (p. Triangularis) |
|  |  | -50 | 18 | 20 |  |
|  |  | -54 | 26 | 22 |  |
|  |  | -54 | 20 | 24 |  |
|  |  | -58 | -44 | 4 | L Middle Temporal Gyrus |
|  |  | -56 | -30 | -6 |  |
|  |  | -58 | -40 | 0 |  |
|  |  | -56 | -50 | 2 |  |
|  |  | -50 | -42 | 4 |  |
|  |  | -42 | 20 | -20 | L Temporal Pole |
| cluster2 | 9581 | 58 | 26 | 0 | R IFG (p. Triangularis) |
|  |  | 54 | 24 | 0 |  |
|  |  | 48 | 30 | 2 |  |
|  |  | 40 | 26 | 12 |  |
|  |  | 52 | 28 | 12 |  |
|  |  | 44 | 26 | -6 | R IFG (p. Orbitalis) |
|  |  | 56 | 30 | -4 |  |
|  |  | 38 | 24 | -10 |  |
|  |  | 36 | 22 | -12 |  |
|  |  | 50 | -32 | 0 | R Middle Temporal Gyrus |
|  |  | 46 | 22 | -4 | R Insula Lobe |
| *Speech fluency* | | | | | |
| cluster1 | 48343 | -40 | 12 | -2 | L Insula Lobe |
|  |  | 40 | 18 | 4 | R Insula Lobe |
|  |  | -6 | 26 | 32 | L MCC |
|  |  | -4 | 14 | 42 |  |
|  |  | 6 | 22 | 38 | R MCC |
|  |  | 2 | 24 | 36 |  |
|  |  | 4 | 28 | 32 |  |
|  |  | 0 | 18 | 40 | L Superior Medial Gyrus |
|  |  | 0 | 14 | 44 | L Posterior-Medial Frontal |
|  |  | -16 | -12 | 6 | L Thalamus |
| cluster2 | 433 | 34 | -62 | -26 | R Cerebelum (VI) |
|  |  | 28 | -62 | -20 |  |
|  |  | 42 | -66 | -22 |  |
|  |  | 34 | -50 | -30 |  |
|  |  | 38 | -48 | -34 | R Cerebelum (Crus 1) |
|  |  | 38 | -62 | -32 |  |
|  |  | 42 | -64 | -26 |  |
|  |  | 38 | -54 | -30 |  |
|  |  | 38 | -62 | -16 | R Fusiform Gyrus |
| cluster3 | 332 | -10 | -92 | 8 | L Superior Occipital Gyrus |
|  |  | -10 | -86 | 6 | L Calcarine Gyrus |
|  |  | -12 | -82 | 2 | L Lingual Gyrus |
|  |  | -22 | -84 | 14 | L Middle Occipital Gyrus |
|  |  | -16 | -100 | 0 |  |
|  |  | -24 | -94 | 8 |  |
|  |  | -22 | -100 | -2 |  |
|  |  | -18 | -88 | -4 |  |
|  |  | -20 | -96 | 0 |  |
| cluster4 | 180 | 26 | -40 | -14 | R Fusiform Gyrus |
|  |  | 34 | -36 | -14 |  |
|  |  | 28 | -36 | -20 |  |
|  |  | 22 | -38 | -18 |  |
|  |  | 16 | -46 | -4 | R Lingual Gyrus |
|  |  | 14 | -42 | 0 |  |
|  |  | 22 | -44 | -8 |  |
|  |  | 18 | -42 | -2 |  |
|  |  | 26 | -42 | -8 | R ParaHippocampal Gyrus |
|  |  | 20 | -42 | -20 | R Cerebelum (IV-V) |
| cluster5 | 176 | -48 | -68 | 24 | L Angular Gyrus |
|  |  | -50 | -68 | 28 |  |
|  |  | -48 | -68 | 32 |  |
|  |  | -50 | -68 | 36 |  |
|  |  | -50 | -68 | 28 |  |
|  |  | -42 | -76 | 28 | L Middle Occipital Gyrus |
| *Task-based category* | | | | | |
| *repetition* | | | | | |
| cluster1 | 46736 | -58 | -40 | 16 | L Superior Temporal Gyrus |
|  |  | 58 | -40 | 22 | R Superior Temporal Gyrus |
|  |  | 54 | -40 | 16 |  |
|  |  | 54 | -40 | 22 |  |
|  |  | 62 | -28 | 14 |  |
|  |  | 62 | -30 | 18 |  |
|  |  | 62 | -34 | 16 |  |
|  |  | 70 | -34 | 18 |  |
|  |  | 68 | -34 | 22 |  |
|  |  | 62 | -36 | 22 |  |
|  |  | 64 | -36 | 26 | R SupraMarginal Gyrus |
| cluster2 | 234 | 4 | -42 | 54 | R Precuneus |
|  |  | 16 | -60 | 44 |  |
|  |  | 6 | -60 | 52 |  |
|  |  | 10 | -46 | 52 |  |
|  |  | 8 | -50 | 58 |  |
|  |  | 8 | -46 | 46 |  |
|  |  | 10 | -60 | 46 |  |
|  |  | 6 | -56 | 56 |  |
|  |  | -6 | -46 | 56 | L Precuneus |
|  |  | -2 | -48 | 54 |  |
|  |  | -2 | -46 | 50 | L MCC |
| cluster3 | 191 | -8 | -64 | 46 | L Precuneus |
|  |  | -12 | -64 | 34 |  |
|  |  | -12 | -68 | 40 |  |
|  |  | -12 | -54 | 38 |  |
|  |  | -10 | -60 | 44 |  |
|  |  | -6 | -60 | 44 |  |
|  |  | -12 | -56 | 34 |  |
|  |  | -10 | -60 | 36 |  |
|  |  | -6 | -64 | 32 |  |
|  |  | -12 | -80 | 36 | L Cuneus |
|  |  | -16 | -74 | 40 | L Superior Parietal Lobule |
| cluster4 | 145 | -30 | -80 | 26 | L Middle Occipital Gyrus |
|  |  | -30 | -80 | 32 |  |
|  |  | -30 | -86 | 30 |  |
|  |  | -30 | -74 | 36 |  |
|  |  | -24 | -80 | 32 | L Superior Occipital Gyrus |
|  |  | -22 | -78 | 36 |  |
| cluster5 | 106 | 36 | 30 | 46 | R Middle Frontal Gyrus |
|  |  | 28 | 34 | 40 |  |
|  |  | 22 | 32 | 42 |  |
|  |  | 36 | 28 | 42 |  |
|  |  | 32 | 32 | 46 |  |
|  |  | 32 | 28 | 46 |  |
|  |  | 38 | 22 | 44 |  |
|  |  | 38 | 26 | 48 |  |
|  |  | 38 | 24 | 40 |  |
|  |  | 32 | 30 | 40 |  |
|  |  | 20 | 32 | 38 | R Superior Frontal Gyrus |
| cluster6 | 102 | 0 | 56 | 6 | L Superior Medial Gyrus |
|  |  | 0 | 58 | 2 |  |
|  |  | 0 | 62 | 6 |  |
|  |  | 4 | 62 | 2 | R Superior Medial Gyrus |
|  |  | 4 | 50 | 6 |  |
|  |  | 8 | 58 | 4 |  |
|  |  | 4 | 58 | -2 | R Mid Orbital Gyrus |
| *naming* | | | | | |
| cluster1 | 9767 | -60 | -12 | 0 | L Superior Temporal Gyrus |
|  |  | -66 | -22 | 6 |  |
|  |  | -62 | -22 | 10 |  |
|  |  | -50 | -14 | 2 |  |
|  |  | -50 | -18 | 6 |  |
|  |  | -62 | -28 | 4 |  |
|  |  | -58 | 0 | -4 |  |
|  |  | -66 | -30 | 4 | L Middle Temporal Gyrus |
|  |  | -64 | -38 | 6 |  |
|  |  | -60 | -8 | 20 | L Postcentral Gyrus |
|  |  | -36 | -30 | 14 | L Heschls Gyrus |
| cluster2 | 8557 | 66 | -12 | 6 | R Superior Temporal Gyrus |
|  |  | 64 | -18 | 8 |  |
|  |  | 62 | -6 | 4 |  |
|  |  | 62 | 0 | 0 |  |
|  |  | 66 | -2 | -4 |  |
|  |  | 62 | -10 | 6 |  |
|  |  | 58 | -10 | 2 |  |
|  |  | 68 | -22 | 4 |  |
|  |  | 62 | -6 | 8 | R Rolandic Operculum |
|  |  | 58 | -10 | 8 | R Heschls Gyrus |
|  |  | 60 | 4 | -4 | R Temporal Pole |
| cluster3 | 1496 | -14 | -60 | -14 | L Cerebelum (VI) |
|  |  | 22 | -54 | 2 | R Lingual Gyrus |
|  |  | 26 | -52 | -10 |  |
|  |  | 24 | -48 | -4 |  |
|  |  | 26 | -54 | 2 |  |
|  |  | 16 | -62 | -2 |  |
|  |  | -6 | -72 | 18 | L Calcarine Gyrus |
|  |  | -10 | -62 | 8 |  |
|  |  | -10 | -64 | 4 |  |
|  |  | -2 | -72 | 20 |  |
|  |  | 4 | -66 | 18 | R Calcarine Gyrus |
| cluster4 | 1420 | 8 | -4 | 56 | R Posterior-Medial Frontal gyrus |
|  |  | 2 | -2 | 62 |  |
|  |  | 8 | -4 | 62 |  |
|  |  | 2 | -2 | 66 |  |
|  |  | 4 | -6 | 56 |  |
|  |  | 2 | -4 | 50 |  |
|  |  | 14 | 0 | 66 |  |
|  |  | -4 | 0 | 60 | L Posterior-Medial Frontal gyrus |
|  |  | -4 | 0 | 66 |  |
|  |  | -2 | -4 | 44 | L MCC |
|  |  | -6 | 12 | 36 |  |
| cluster5 | 741 | -16 | -24 | 6 | L Thalamus |
|  |  | -12 | -28 | -2 |  |
|  |  | -20 | -24 | 8 |  |
|  |  | -16 | -24 | 0 |  |
|  |  | -18 | -20 | 6 |  |
|  |  | -10 | -24 | 0 |  |
|  |  | -4 | -24 | 2 |  |
|  |  | -10 | -28 | 2 |  |
|  |  | -26 | -50 | -2 | L Lingual Gyrus |
|  |  | -24 | -46 | -8 |  |
|  |  | -14 | -28 | -10 | L Hippocampus |
| cluster6 | 200 | 22 | 28 | -14 | R Superior Orbital Gyrus |
|  |  | 28 | 32 | -18 | R IFG (p. Orbitalis) |
|  |  | 28 | 28 | -14 |  |
|  |  | 38 | 38 | -12 |  |
|  |  | 44 | 32 | -4 |  |
|  |  | 46 | 36 | -14 |  |
|  |  | 44 | 34 | -8 |  |
|  |  | 40 | 42 | -10 |  |
|  |  | 34 | 36 | -12 |  |
|  |  | 38 | 30 | -8 |  |
| cluster7 | 194 | -14 | -12 | 20 | L Caudate Nucleus |
|  |  | -14 | -8 | 22 |  |
|  |  | -12 | -6 | 18 |  |
|  |  | -6 | -10 | 10 | L Thalamus |
| cluster8 | 148 | -8 | -38 | 46 | L MCC |
|  |  | -10 | -34 | 44 |  |
|  |  | -6 | -34 | 44 |  |
|  |  | -10 | -42 | 42 |  |
|  |  | -4 | -42 | 46 |  |
|  |  | -12 | -34 | 40 |  |
|  |  | -14 | -38 | 44 |  |
|  |  | -8 | -30 | 44 |  |
|  |  | -6 | -28 | 40 |  |
|  |  | -6 | -34 | 36 |  |
| cluster9 | 138 | -36 | -32 | -14 | L Inferior Temporal Gyrus |
|  |  | -34 | -32 | -18 | L Fusiform Gyrus |
|  |  | -28 | -36 | -20 |  |
|  |  | -34 | -36 | -26 |  |
|  |  | -34 | -30 | -22 |  |
|  |  | -32 | -34 | -10 | L Hippocampus |
|  |  | -34 | -28 | -14 |  |
|  |  | -34 | -30 | -10 |  |
|  |  | -28 | -34 | -14 | L ParaHippocampal Gyrus |
| cluster10 | 111 | -2 | -44 | 20 | L PCC |
|  |  | 6 | -42 | 18 | R PCC |
|  |  | 10 | -44 | 20 |  |
|  |  | 12 | -44 | 28 |  |
|  |  | 10 | -40 | 26 |  |
|  |  | 6 | -44 | 26 |  |
| cluster11 | 106 | 16 | -72 | 32 | R Cuneus |
|  |  | 14 | -72 | 38 |  |
|  |  | 14 | -68 | 40 | R Precuneus |
| cluster12 | 100 | -46 | 46 | -10 | L Middle Orbital Gyrus |
|  |  | -42 | 44 | -2 |  |
|  |  | -46 | 48 | -6 |  |
|  |  | -42 | 42 | -16 | L IFG (p. Orbitalis) |
|  |  | -46 | 44 | -6 |  |
| *reading* | | | | | |
| cluster1 | 32973 | -42 | 22 | -8 | L IFG (p. Orbitalis) |
|  |  | -50 | 22 | 10 | L IFG (p. Triangularis) |
|  |  | -46 | 22 | 14 |  |
|  |  | -50 | 16 | 22 | L IFG (p. Opercularis) |
|  |  | -54 | 16 | 20 |  |
|  |  | -58 | -34 | 0 | L Middle Temporal Gyrus |
|  |  | -58 | -36 | 4 |  |
|  |  | -58 | -40 | 2 |  |
|  |  | -54 | -44 | 2 |  |
|  |  | -58 | -44 | 4 |  |
|  |  | -56 | -50 | 4 |  |
| cluster2 | 2359 | -6 | -66 | 8 | L Lingual Gyrus |
|  |  | 6 | -66 | 4 | R Lingual Gyrus |
|  |  | -8 | -70 | 8 | L Calcarine Gyrus |
|  |  | -10 | -64 | 8 |  |
|  |  | -4 | -56 | 36 | L Precuneus |
|  |  | 0 | -50 | 40 |  |
|  |  | -4 | -50 | 14 |  |
|  |  | -6 | -52 | 40 |  |
|  |  | 8 | -52 | 14 | R Precuneus |
|  |  | 16 | -78 | -30 | R Cerebelum (Crus 1) |
| cluster3 | 575 | 50 | -62 | 18 | R Middle Temporal Gyrus |
|  |  | 52 | -50 | 20 |  |
|  |  | 58 | -52 | 16 |  |
|  |  | 50 | -58 | 20 |  |
|  |  | 48 | -58 | 16 |  |
|  |  | 52 | -58 | 10 |  |
|  |  | 56 | -50 | 22 | R Superior Temporal Gyrus |
|  |  | 44 | -50 | 22 | R Angular Gyrus |
|  |  | 48 | -66 | 34 |  |
|  |  | 44 | -70 | 40 |  |
|  |  | 48 | -44 | 24 | R SupraMarginal Gyrus |
| cluster4 | 304 | 30 | -74 | -30 | R Cerebelum (Crus 1) |
|  |  | 32 | -50 | -30 | R Cerebelum (VI) |
|  |  | 36 | -66 | -24 |  |
|  |  | 30 | -66 | -22 |  |
|  |  | 40 | -40 | -24 | R Fusiform Gyrus |
|  |  | 32 | -44 | -20 |  |
|  |  | 42 | -48 | -22 |  |
|  |  | 38 | -34 | -18 |  |
|  |  | 32 | -40 | -18 |  |
|  |  | 40 | -40 | -18 |  |
|  |  | 46 | -48 | -20 | R Inferior Temporal Gyrus |
| cluster5 | 167 | 38 | -76 | -4 | R Inferior Occipital Gyrus |
|  |  | 40 | -78 | -8 |  |
|  |  | 36 | -70 | -10 |  |
|  |  | 36 | -74 | -12 | R Fusiform Gyrus |
|  |  | 30 | -70 | -14 |  |
|  |  | 36 | -82 | 2 | R Middle Occipital Gyrus |
|  |  | 40 | -78 | 4 |  |
|  |  | 36 | -84 | 10 |  |
| cluster6 | 138 | 32 | -76 | 32 | R Middle Occipital Gyrus |
|  |  | 32 | -76 | 40 |  |
|  |  | 28 | -76 | 26 |  |
|  |  | 34 | -70 | 30 |  |
|  |  | 28 | -78 | 30 | R Superior Occipital Gyrus |
|  |  | 28 | -76 | 38 |  |
|  |  | 26 | -74 | 32 |  |
|  |  | 20 | -70 | 38 |  |
|  |  | 22 | -76 | 38 |  |
|  |  | 26 | -70 | 38 |  |
|  |  | 28 | -70 | 26 |  |
| cluster7 | 101 | 40 | -26 | 46 | R Postcentral Gyrus |
|  |  | 36 | -34 | 44 |  |
|  |  | 34 | -32 | 48 |  |
|  |  | 44 | -24 | 50 |  |
|  |  | 44 | -24 | 56 |  |
|  |  | 34 | -30 | 44 |  |
|  |  | 46 | -18 | 52 |  |
|  |  | 40 | -38 | 48 | R Inferior Parietal Lobule |
|  |  | 40 | -32 | 42 | R SupraMarginal Gyrus |
|  |  | 40 | -34 | 46 |  |
| cluster8 | 100 | -62 | -30 | 36 | L SupraMarginal Gyrus |

**Table S1**. The results of Neurosynth meta-analytic coactivation maps of the ALE clusters.

**Supplementary Table 2**

|  | category | keywords |
| --- | --- | --- |
| Language  function | Semantics | semantic, sentences, retrieved, recall, read, written, word, comprehension, lexical, language |
|  | Phonology | auditory, speech, speech production, acoustic, speech perception, listening, sounds, audiovisual, orthographic, silent, musical, reading |
|  | Speech fluency | pain, phonological, auditory, speech, acoustic, speech perception, sounds, audiovisual, speech production |
| Task | Repetition | auditory, motor, syntactic, verbs, hearing, production, phonological, speech production, video, mirror |
|  | Reading | auditory, sounds, speech, listening, acoustic, language, comprehension, semantic, words, sentences |
|  | Naming | episodic, recall, autobiographical, balance, amnestic, imagine, semantic memory, auditory, speech, acoustic |

Table S2. Keywords from each functional/task-based categories
